# Evaluation of the antibody response and adverse reactions of the BNT162b2 vaccine of participants with prior COVID-19 infection in Japan

**DOI:** 10.1101/2021.07.18.21260579

**Authors:** Toshiya Mitsunaga, Yuhei Ohtaki, Yutaka Seki, Masakata Yoshioka, Hiroshi Mori, Midori Suzuka, Syunsuke Mashiko, Satoshi Takeda, Kunihiro Mashiko

**Author notes:** Corresponding Author’s Information: Name: Toshiya Mitsunaga, Address: 3-10-1, Sanda-machi, Hachioji-city, Tokyo, 1930832, Japan.

## Abstract

**Introduction:** Vaccination programs are important to preventing COVID-19 infection. BNT162b2 is new type of vaccine, and previous studies have shown that the antibody response was significantly elevated in patients with prior COVID-19 infection after the first vaccination. However, no study has evaluated the efficacy of the vaccination or the adverse reactions of people with prior COVID-19 infection in Japan. The aim of this study is to evaluate the antibody titer and adverse reactions of BNT162b2 vaccine among participants with prior COVID-19 infection in Japan.

**Methods:** The data for this study was collected between April 15, 2021, and June 9, 2021. All of the hospital staff who received the BNT162b2 vaccine were included in this study and were sorted into either the prior infection group or the control group. We collected the data of adverse reactions through self-reporting and calculated the anti-SARS-CoV-2 spike-specific antibody titer for all participants.

**Results:** The antibody titer of the prior-infection group in first antibody test was significantly higher than that of the control group in the second antibody test. There was no significant difference in adverse reactions between the prior infection group receiving its first vaccination and the control group receiving its second vaccination.

**Conclusion:** Our study shows that the antibody response following the first vaccination in the prior COVID-19 infection group was found to be comparable to that of the second vaccination in the control group; however, the evaluation of adverse reactions was inadequate and further, large-scale studies are needed.

## Introduction

At the end of 2019, a new pathogen named the severe acute respiratory syndrome coronavirus 2 (SARS-CoV-2) was identified. The coronavirus 2019 (COVID-19) has since spread rapidly all over the world, and the number of COVID-19 cases has reached more than 180 million with four million COVID-19 patients dead (mortality rate: 2.16%) [1]. In Japan, The Ministry of Health, Labor and Welfare has reported that the number of COVID-19 cases in Japan was 798,159, with 14,740 deaths by 30 June 2021 (mortality rate: 1.85%) [2]. Countries around the world are trying to reduce their rates of COVID-19 infection, but this is difficult to control due to the virus’s strong infectivity. Treatment plans such as steroids, antivirals, ventilators, etc. have been put in place, but vaccination is still a crucial strategy for preventing COVID-19 infection. BNT162b2 is a new type of vaccine composed of a nucleoside-modified RNA vaccine that encodes a full-length SARS-CoV-2 spike glycoprotein [3]. A study carried out by Polack et al. showed that the vaccine’s efficacy for preventing COVID-19 is about 95% [3], and we are administering two doses of the vaccine to boost immunity. The study carried by Walsh et al. showed that spike-binding IgG titer increased dramatically and reached a plateau after seven days of receiving the second vaccination, whereas the titer was slightly elevated after seven days of receiving the first vaccination [4].

Previous studies carried out by Charlotte et al. and Ebinger et al. showed that the antibody-titer level was significantly elevated in patients with a history of prior COVID-19 infection after receiving the first vaccination [5, 6] but this level was merely maintained after these patients received the second vaccination. Although there are several studies in this field, no study has evaluated the effects of the first and second vaccinations on people who had previously experience a COVID-19 infection in Japan. Furthermore, no study has evaluated the adverse reactions to the BNT162b2 vaccine for people with prior COVID-19 infection. The aim of this study is to evaluate the antibody titer and adverse reactions to the BNT162b2 vaccine among subjects with a history of COVID-19 infection in Japan.

## Materials and Methods

### Study Design and Setting

This prospective study, which builds upon our previous study [7] by adding the data of a prior COVID-19 infection group, was carried out between April 15, 2021, and June 9, 2021, at the Association of EISEIKAI Medical and Healthcare Corporation Minamitama Hospital, an urban hospital with 170 beds. The protocol for this research project was approved by the Ethics Committee of the institution, it conforms to the provision of the Declaration of Helsinki (Committee of the Association of EISEIKAI Medical and Healthcare Corporation Minamitama Hospital, Approval No. 2020-Ack-19) and written consent was obtained from all the participants.

All our hospital’s staff who received the BNT162b2 vaccine as a part of a routine program were included in this study. The exclusion criteria were as follows: 1) cases did not give informed consent, 2) cases that exhibited a new COVID-19 infection following vaccination, 3) cases that did not provide a blood sample within the expected deadline (i.e., twenty days post-vaccination), and 4) cases that did not submit a questionnaire about adverse reactions.

The participants with a history of prior COVID-19 infection or whose antibody titer was positive before the administration of the BNT162b2 vaccine were included in this study as part of the prior COVID-19 infection group, whereas the participants without a history of prior COVID-19 infection or who presented with a negative antibody titer before vaccination were included as the control group.

We obtained 5-ml blood samples from the intermediate cubital vein and calculated the anti-SARS-CoV-2 spike-specific antibody titer for each participant (Elecsys^®^ Anti-SARS-CoV-2 S RUO, Roche Diagnostics K.K.) 1) before vaccination (baseline), 2) seven to twenty days after the first dose of the vaccination (first antibody test), and 3) seven to twenty days after the second dose of the vaccination (second antibody test).

We also calculated the blood cell count (WBC: White Blood Cells, Hb: Hemoglobin, Ht: Hematocrit, and PLT: Platelets) and conducted biochemical examinations (AST: aspartate aminotransferase, ALT: alanine aminotransferase, γ-GT: γ-glutamyl transpeptidase, Alb: albumin, TG: triglyceride, HDL-C: high density lipoprotein cholesterol, LDL-C: low density lipoprotein cholesterol, Cr: Creatinine, BS: Blood Sugar, HbA1c: Hemoglobin A1c, CRP: c-reactive protein) before vaccination.

The information of age; gender; height; body weight; body mass index (BMI); past medical history (1. Chronic lung disease (Chronic obstructive pulmonary disease: COPD), 2. Chronic lung disease (non-COPD), 3. Cardiac disease, 4. Hypertension, 5. Diabetes, 6. Dyslipidemia, 7. Liver disease, 8. Chronic kidney disease, 9. Autoimmune disease, 10. Cancer); medication (1. Antihypertensive drugs, 2. Antidiabetic drugs, 3. Antilipid drugs, 4. Antiplatelet and Anticoagulant drugs, 5. Immunosuppressive drugs, 6. Immunoglobulin); smoking habits (current and previous history of smoking); habits of drinking alcohol; and adverse reactions (1. pain/redness/swelling, 2. fever, 3. fatigue, 4. headache, 5. chills, 6. diarrhea, 7. myalgia, 8. arthralgia, 9. others) were recorded via self reporting. Cases with missing values were not excluded, and the acquired values were used for analysis.

### Statistical Analyses

A sample size of 475 participants was determined based upon 70% power, 0.05 significance level, 0.8 effect size, 37 allocation ration, and 20% attrition.

Continuous variables were described as medians and interquartile ranges (IQR) and were compared using a Mann-Whitney *U*-test. Categorical variables were expressed as numbers and percentages and were compared using Fisher’s exact test. Analysis of variance (ANOVA) and Fisher’s exact test were used to test differences among the groups. Multivariate logistic regression analysis was performed to ascertain the effects of factors such as the likelihood of local adverse reactions or systemic adverse reactions. Odds ratios and corresponding 95% confidence intervals were calculated. A *p*-value of less than 0.05 was considered to indicate statistical significance. Data were analyzed with the Statistical Package for the Social Sciences, version 26.0 (SPSS, Chicago, IL, USA).

## Results

Overall, 501 participants met the inclusion criteria. However, 96 participants were excluded because they did not provide their consent to the study, one participant was excluded because of a new COVID-19 infection after vaccination, and twenty-one participants were excluded because they did not provide a blood sample within the deadline. Finally, 383 (76.4%) participants, who were divided into a nine-person prior COVID-19 infection group and a 374-person control group, were analyzed (Fig 1).

**Fig.1.**
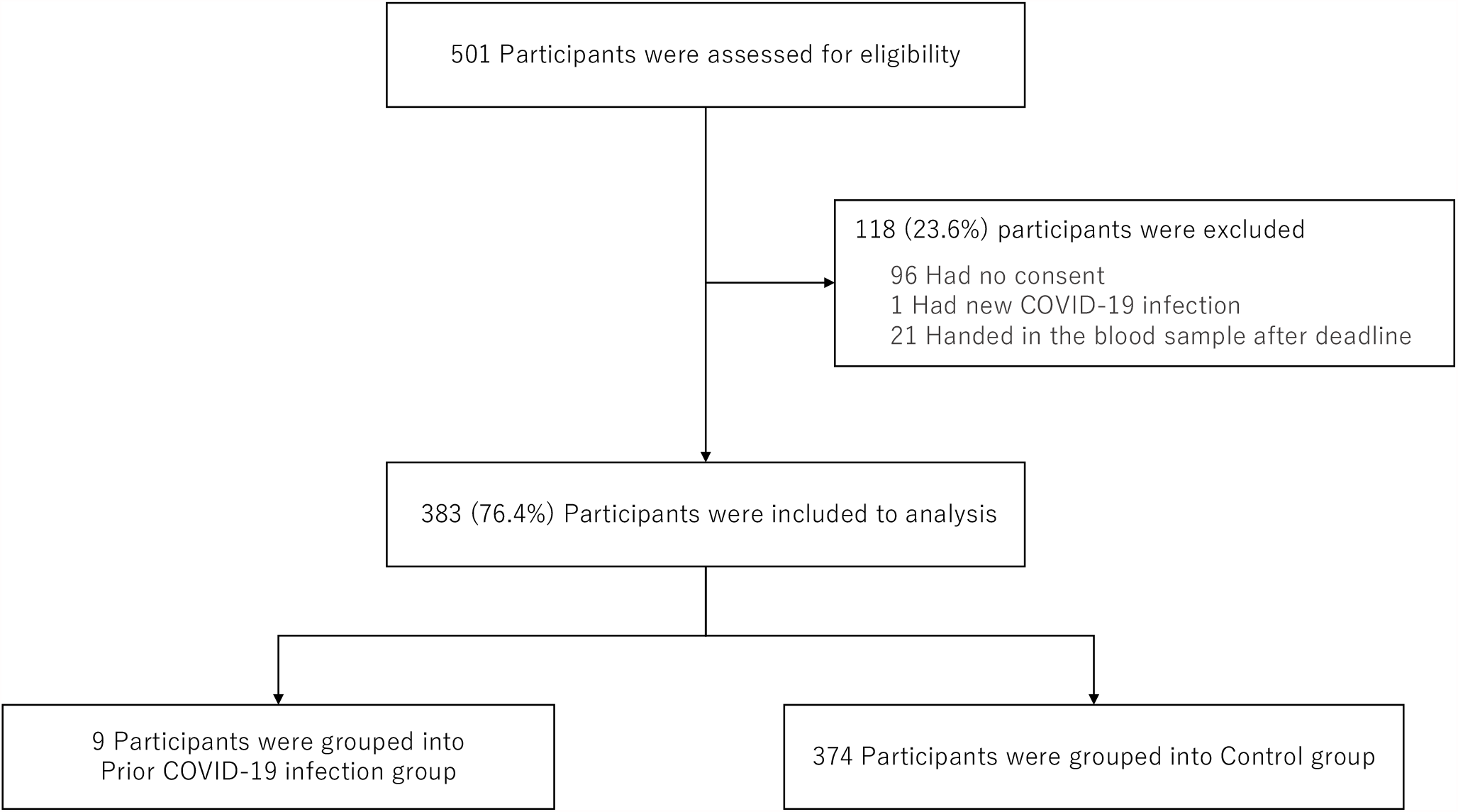
Flow chart of participant’s selection with inclusion and exclusion data

The data of the antibody titer of the second antibody test in the prior COVID-19 infection group was missing for one case, and the data of adverse reactions was missing for two cases in the prior COVID-19 infection group and for fifteen cases in control group.

The baseline characteristics for the prior COVID-19 infection group and control group are demonstrated in Table 1. The median age (interquartile range) of the participants was younger in the prior COVID-19 infection Group than in the Control Group (26 (16.0) vs 36 (16.0): *p* = 0.12), but there were no other significant differences. The proportion of males was slightly smaller in the prior COVID-19 infection group, but the difference was not significant (*p* = 0.46). Moreover, there was no obesity and only a few participants with a previous medical history in the prior COVID-19 infection group.

**Table 1.**
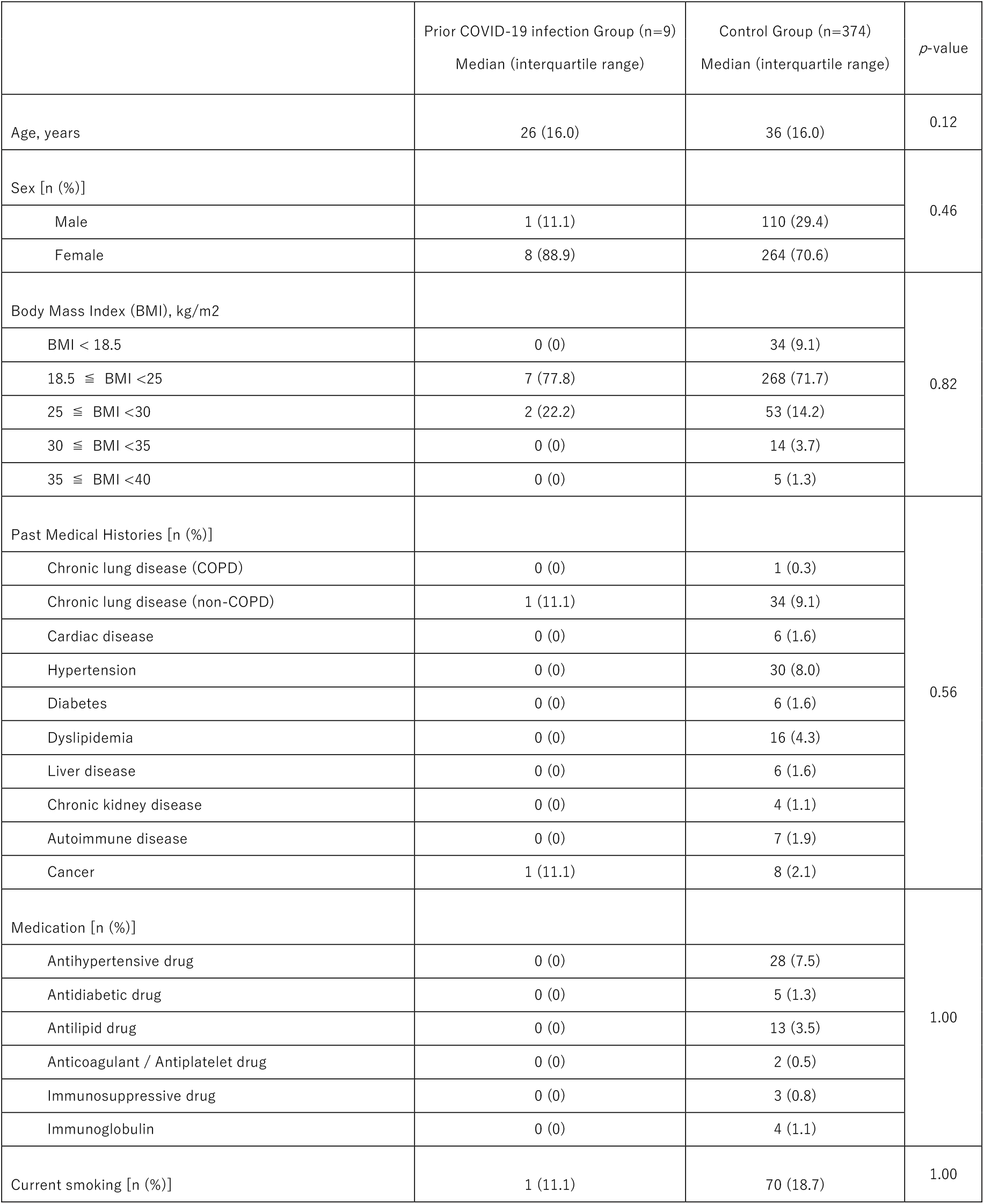
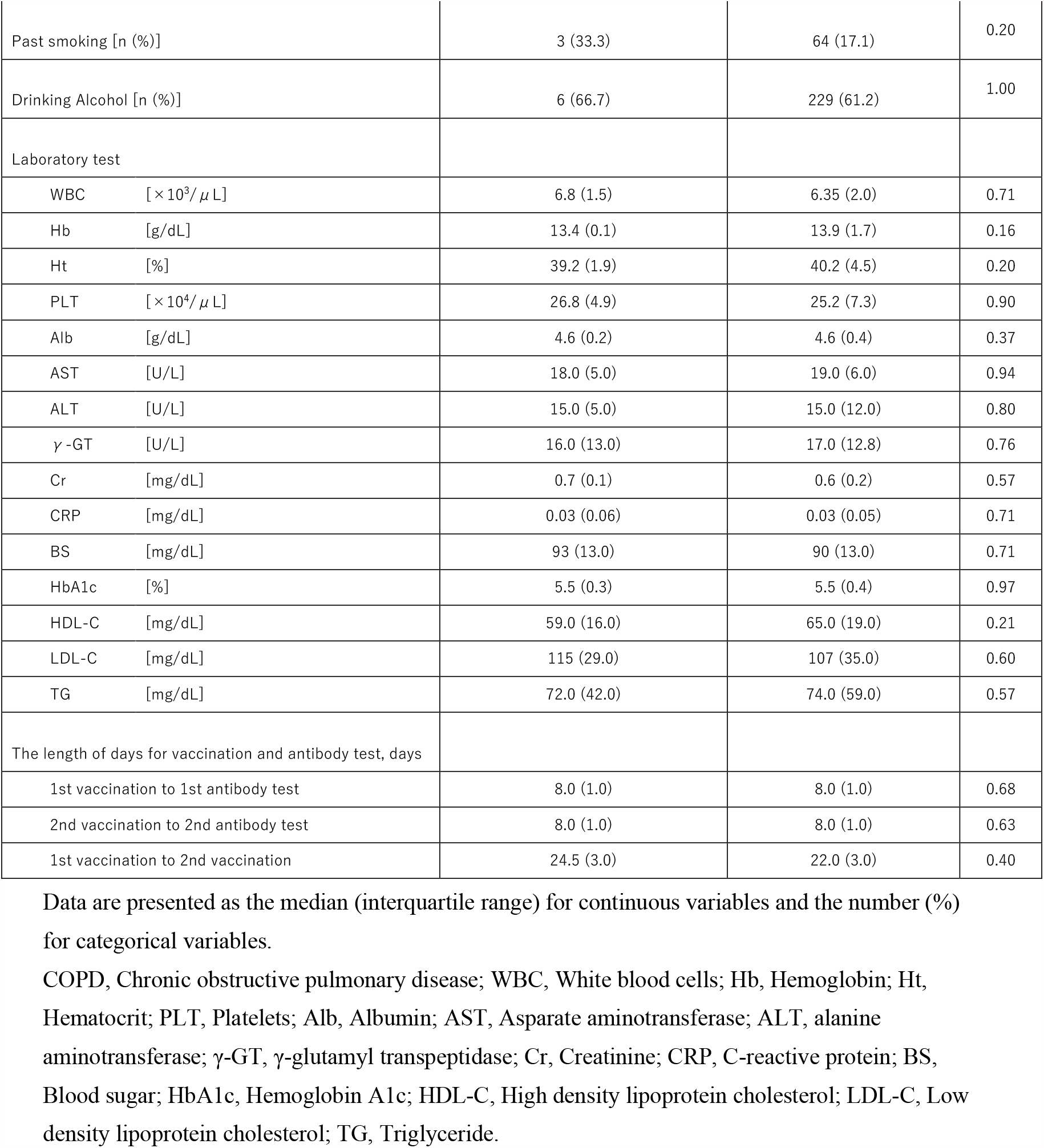
Baseline characteristics of the study population.

The comparison of anti-SARS-CoV-2 spike-specific antibody response between the prior COVID-19 infection group and the control group is demonstrated in Fig 2. The log anti-SARS-CoV-2 spike-specific antibody titers of the prior COVID-19 infection group were higher than that of the control group in the pre-vaccination, first antibody, and second antibody tests (*p* < 0.001). The log antibody titer of the prior COVID-19 infection group in the first antibody test was significantly higher than that of control group in second antibody test (*p* < 0.001), whereas there was no significant difference of the log antibody titer of the prior COVID-19 infection group between the first antibody test and the second antibody test (*p* = 0.54).

**Fig. 2.**
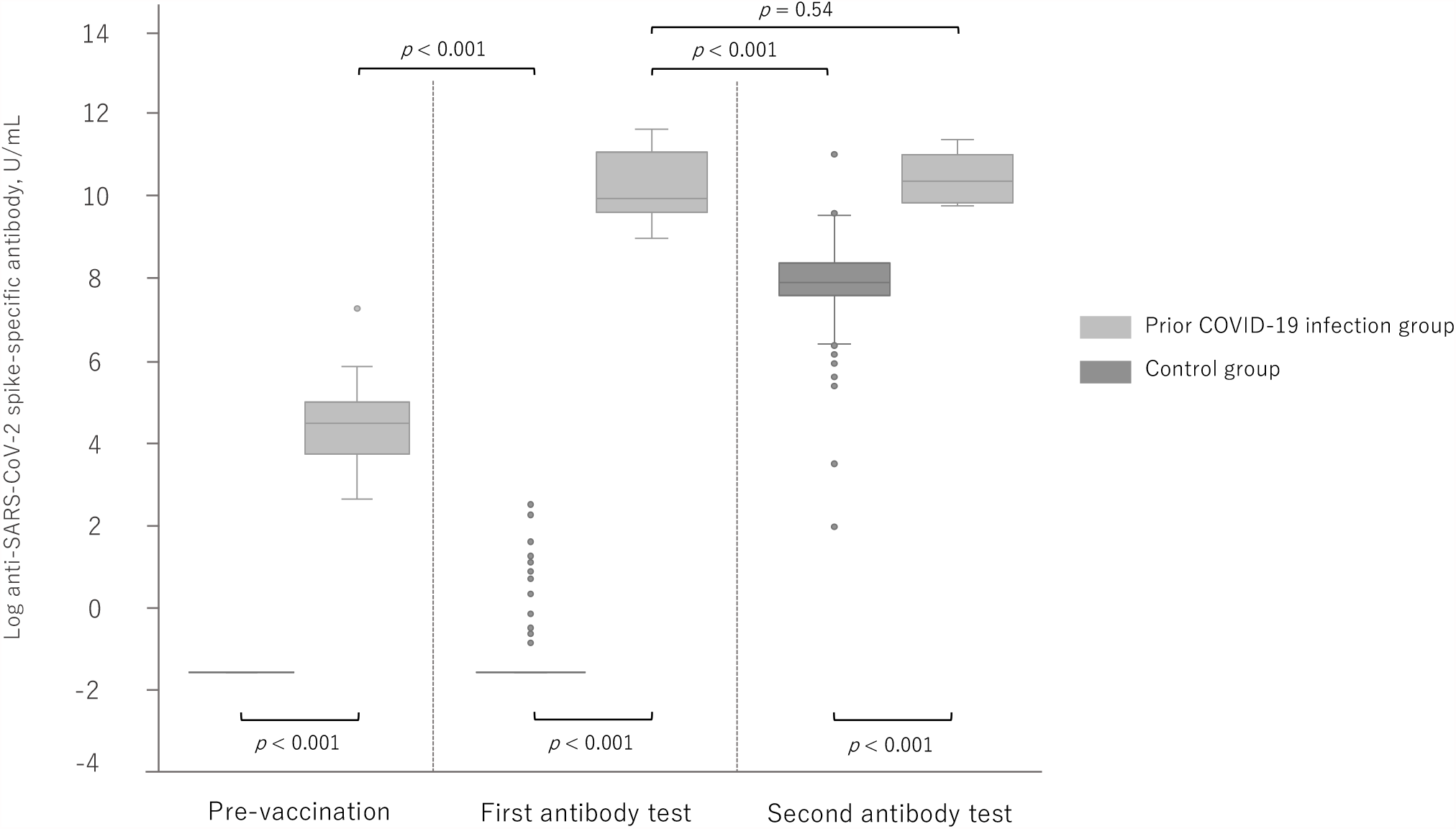
Anti-SARS-CoV-2 spike-specific antibody response after the administration of the BNT162b2 vaccine to participants with and without prior COVID-19 infection.

The number of adverse reactions after the administration of BNT162b2 vaccine is demonstrated in Table 2. In the prior COVID-19 infection group, the proportion of symptoms was not different between the first and second vaccinations. There was no significant difference of adverse reaction between the first vaccination in the prior COVID-19 infection group and in the second vaccination in control group (*p* = 0.84). The proportion of systemic symptoms of second vaccination in each group was larger than that of the first vaccination in the control group (*p* < 0.001).

**Table 2.** The number of adverse reactions after the administration of the BNT162b2 vaccine between the two groups.

|  | Prior COVID-19 infection Group (n=7) |  | Control Group (n=359) |  | p-value |  |  |  |  |  |
| --- | --- | --- | --- | --- | --- | --- | --- | --- | --- | --- |
|  | First vaccination<br>(pFV) | Second vaccination<br>(pSV) | First vaccination<br>(cFV) | Second vaccination<br>(cSV) | pFV vs<br>pSV | pFV vs<br>cFV | pFV vs<br>cSV | pSV vs<br>cFV | pSV vs<br>cSV | cFV vs<br>cSV |
| The number of the cases (%) |  |  |  |  |  |  |  |  |  |  |
| Injection site symptoms | 6 (85.7) | 6 (85.7) | 352 (98.1) | 351 (97.8) | 0.64 | 0.24 | 0.84 | $p < 0.001$ | 0.91 | $p < 0.001$ |
| fever | 1 (14.3) | 5 (71.4) | 5 (1.4) | 161 (44.8) |  |  |  |  |  |  |
| fatigue | 2 (28.6) | 4 (57.1) | 99 (27.6) | 249 (69.4) |  |  |  |  |  |  |
| headache | 2 (28.6) | 3 (42.9) | 77 (21.4) | 215 (59.9) |  |  |  |  |  |  |
| chills | 0 (0) | 3 (42.9) | 23 (6.4) | 141 (39.3) |  |  |  |  |  |  |
| diarrhea | 0 (0) | 0 (0) | 27 (7.5) | 50 (13.9) |  |  |  |  |  |  |
| myalgia | 3 (42.9) | 2 (28.6) | 106 (29.5) | 166 (46.2) |  |  |  |  |  |  |
| arthralgia | 2 (28.6) | 3 (42.9) | 27 (7.5) | 160 (44.6) |  |  |  |  |  |  |
| others | 0 (0) | 0 (0) | 38 (10.6) | 71 (19.8) |  |  |  |  |  |  |

The proportion of adverse reactions for each day after vaccination is demonstrated in Fig 3. The injection site symptoms and headaches were confirmed in both groups four days after the first and second vaccinations. Fever was confirmed in the early days after the second vaccination in both groups. Fatigue was confirmed after four days of the first and second vaccination doses in the control group, whereas that was confirmed in the early days after both doses in the prior COVID-19 infection group. Myalgia and arthralgia were confirmed after four days of the first and second vaccination in the control group, whereas they were confirmed only within three days after receiving the second dose of the vaccination in the prior COVID-19 infection group.

**Fig. 3.**
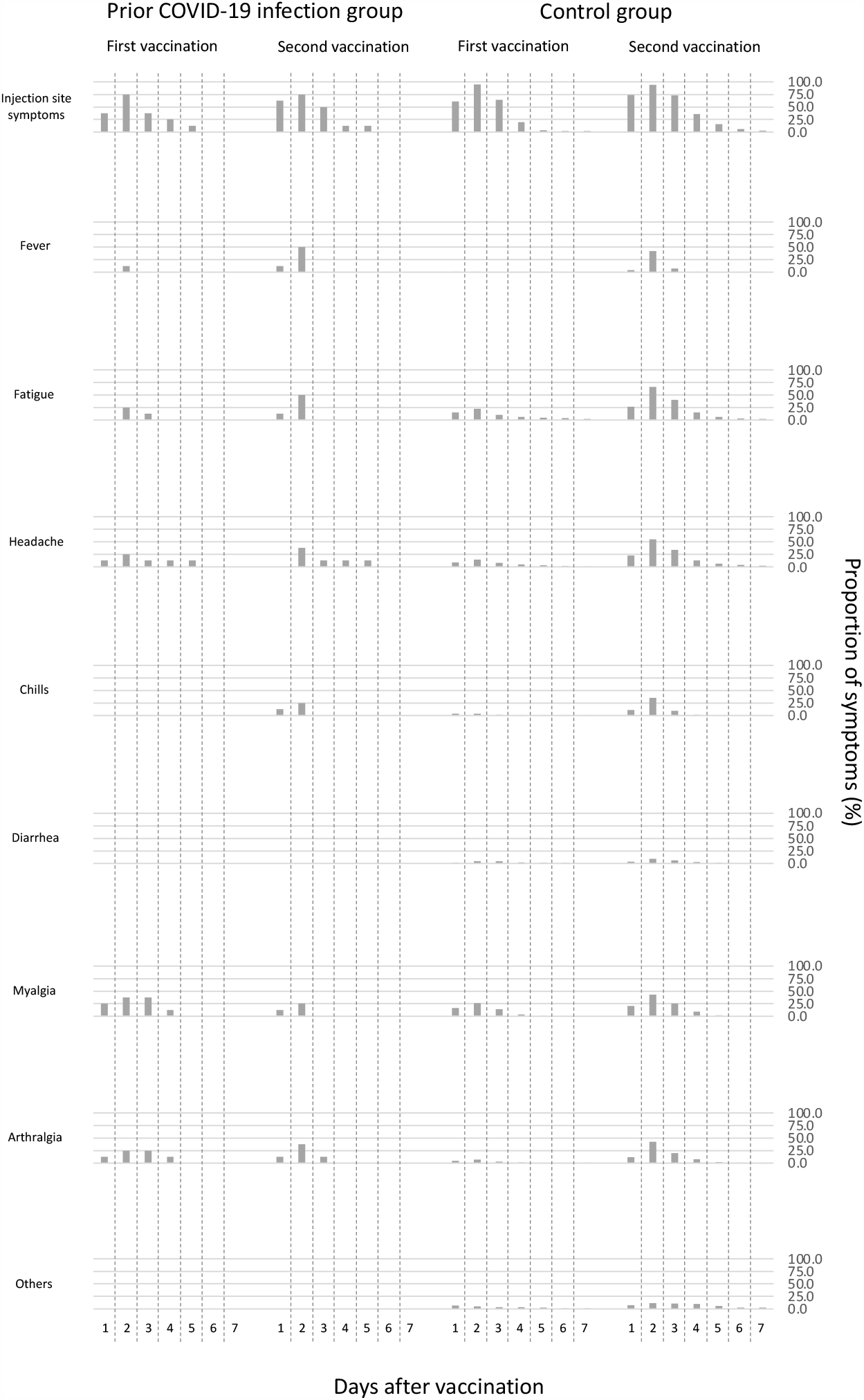
The proportion of adverse reactions for each day after the administration of the BNT162b2 vaccine.

The total days of adverse reactions is recorded in Fig 4. The length of the days of systemic symptoms was approximately half the length of the local symptoms, and the mean of days was shorter than 1.5 days. The total days of fever and fatigue after the second vaccination was significantly longer than after the first vaccination in both groups (*p* < 0.05, *p* < 0.001). In the control group, the total number of days of myalgia and arthralgia was significantly longer after the second dose of the vaccination than that after the first vaccination (*p* < 0.001), whereas the total number of days of myalgia and arthralgia was longer after the first vaccination than after the second vaccination in the prior COVID-19 infection group.

**Fig. 4.**
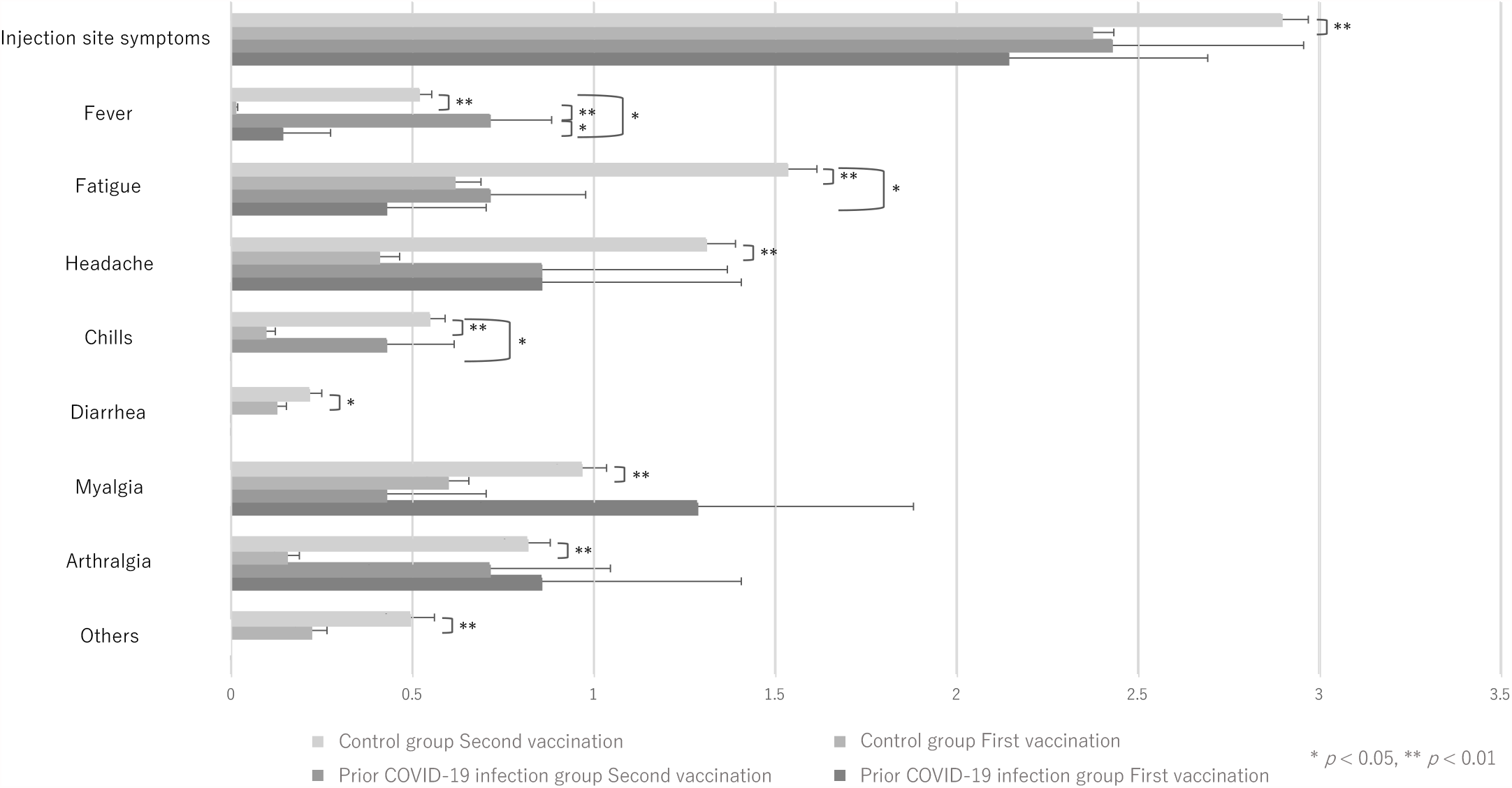
The total days of adverse reactions after the administration of the BNT162b2 vaccine.

Table 3 shows the multivariate logistic regression analysis of the factors associated with local adverse reactions or systemic adverse reactions after receiving the first vaccination. There were no factors affecting local adverse reactions, whereas females and younger participants experienced significantly enhanced systemic adverse reactions (*p* < 0.001, *p* < 0.01). The history of prior COVID-19 infections was not related to local and systemic adverse reactions (*p* = 0.15, *p* = 0.38).

**Table 3.** The multivariate logistic regression analysis of the factors associated with local and systemic adverse reactions after receiving the first vaccination.

| Symptoms/Predictor | Adjusted OR (95% CI) | *p*-value |
| --- | --- | --- |
| Local reactions |  |  |
| Female | 0.47 (0.05 – 4.19) | 0.50 |
| Age | 1.04 (0.97 – 1.14) | 0.26 |
| Body Mass Index: BMI | 1.03 (0.83 – 1.29) | 0.77 |
| Prior Covid-19 infection | 0.18 (0.02 – 1.82) | 0.15 |
| Systemic reactions |  |  |
| Female | 4.12 (2.45 – 6.90) | *p* < 0.001 |
| Age | 0.97 (0.95 – 0.99) | *p* < 0.01 |
| Body Mass Index: BMI | 1.05 (0.99 – 1.12) | 0.13 |
| Prior Covid-19 infection | 0.50 (0.11 – 2.36) | 0.38 |

## Discussion

In our study, the antibody titer dramatically increased in the prior COVID-19 infection group after first vaccination among Japanese people. This is the first study that evaluated the antibody response after administration of the BNT162b2 vaccine in the prior COVID-19 infection group in Japan.

According to previous studies, the median antibody titers after the first dose of vaccination was significantly higher in participants who had been previously infected with the COVID-19 virus than that in uninfected participants [5, 6]. A study carried out by Ebinger et al. shows that the antibody titer reached a plateau after the first dose of the vaccination and that there was no significant difference of antibody titers after the first dose of the vaccination and after the second dose of the vaccination [6]. In our study, the antibody titer of the prior COVID-19 infection group reached a plateau after receiving first dose of the vaccination, and it was significantly higher than that of the control group after receiving the second dose of the vaccination as demonstrated in previous studies. In contrast to the results of previous studies, in our study, the antibody titer of the prior COVID-19 infection group after receiving the first dose of the vaccination was significantly higher than that of the control group after receiving the second dose of the vaccination.

Based on the results of previous studies, age and gender can be considered to be factors that increased the antibody titer. A study carried by Gustafson et al. showed that the immune response to vaccination is controlled by a delicate balance of effector T cells and follicular T cells and aging disturbs this balance. Multiple changes in T cells have been identified as contributing to the age-related defects of post-transcriptional regulation, metabolic function, and T-cell receptor signaling [8]. In this study, there was no significant difference in age between the prior COVID-19 infection group and the control group, but the median age was higher in the prior COVID-19 infection group, which may have increased the antibody titer.

Differences in sex hormones are associated with gender differences in vaccine-induced immunity. For example, testosterone levels and the antibody titer of the influenza vaccine have been shown to be inversely correlated [9–11]. Genetic differences, as well as sex hormone differences, affect vaccine-induced immunity. The X-chromosome expresses ten times more genes than the Y-chromosome, and the differences in gene expression between the X- and Y-chromosomes promote differences in vaccine-induced immunity by gender [12]. In contrast to this widely held belief, in our previous study, gender was not a significant factor in the differences in antibody titers, and it is possible that gender differences did not contribute much to the increases in antibody titers in this study as well [7].

The antibody titer level does not necessarily reflect the immune function against the BNT162b2 vaccine as a whole. However, our study showed that prior COVID-19 infection may increase the immune response after receiving the first dose of the vaccination, and this fact is especially important for vaccine delivery systems in Japan.

There are very few studies that evaluate the adverse reactions of the BNT162b2 vaccine. The proportion of adverse reactions to the first and second doses of the vaccination in the control group was similar to previous studies [4, 13]. Few studies have shown the adverse reactions to receiving the first and second doses of the vaccination with and without prior COVID-19 infection [6, 14, 15], and they have described the number of days and cases after vaccination, but the range of days were two days or fewer, two to seven days, and seven days or more. On the other hand, the number of adverse reactions were recorded on a daily basis up to the seventh day after vaccination in our study. The trend of fever and fatigue in our study was similar to that of the previous study, whereas injection-site symptoms after the first and second vaccinations in both groups, headache in the prior COVID-19 infection group after both doses, and myalgia and arthralgia after the first vaccination in the prior COVID-19 infection group lasted longer than those of previous study [6].

There was no significant difference in the proportion of adverse reactions between the second vaccination in the control group and the first vaccination in the prior COVID-19 infection group. This fact may indicate that prior COVID-19 infection enhances the adverse reaction. On the other hand, in multivariate logistical regression analysis, the history of prior COVID-19 infection was not related to local and systemic adverse reactions, which is likely to be influenced by the sample size. Most adverse reactions in the prior COVID-19 infection group increased after the second vaccination as compared to the adverse reactions after receiving the first vaccination. However, myalgia or arthralgia were reduced or had their duration shortened after patients received the second vaccination. In other words, in the prior COVID-19 infection group, there may be symptom specificity not only in the frequency of adverse reactions, but also in their timing and duration.

As mentioned above, our study demonstrates the possibility of enhanced adverse reactions after the first vaccination and a further enhanced adverse reaction to the second vaccination. Judging comprehensively based on the results of antibody titers, it should be recognized that in Japan a single administration of the BNT162b2 vaccine may be sufficient for pre-infected individuals. Further large-scale studies are necessary to evaluate the immune response and the adverse reactions in the prior COVID-19 infection group.

Our study has several limitations. First, we did not calculate the lymphocyte activity of either group, and we could not monitor the whole immune system response. Second, although we established a relatively large effect size and although the difference in antibody titer between the two groups was clear, we may have needed a large sample size to assess adverse reactions. Third, we only included relatively healthy medical workers, not participants with severe complications, and this may have reduced the differences between the prior COVID-19 infection group and the control group. In conclusion, our study showed that the antibody response to the first vaccination in the prior COVID-19 infection group was found to be comparable to that of the second vaccination in the non-COVID-19 infection group. On the other hand, the evaluation of adverse reactions is inadequate, and further large-scale studies are needed.

## Data Availability

We permit the use of the data available from the links below.

https://www.amazon.co.jp/clouddrive/share/RojSjHadBhY2LHhheayyqxRKphwwV51IP8tXscuA2E8

## IRB Approval Code and Name of the Institution

Approval No. 2020-Ack-19 / Committee of Association of EISEIKAI Medical and Healthcare Corporation Minamitama Hospital.

## Acknowledgements

none.

